# Racial Differences and Temporal Trends in Parkinson’s and Alzheimer’s Disease-Related

**DOI:** 10.64898/2026.10.22.26366001

**Authors:** Md Roungu Ahmmad

**Affiliations:** Biostatistics and Data Science USF Health College of Nursing University of South Florida Tampa, Florida, United States

**Keywords:** Alzheimer’s disease, Parkinson’s disease, Racial disparities, Multiple-cause mortality, Temporal trends, CDC WONDER

## Abstract

**Objective:** The objectives of this study were to examine racial differences and temporal trends in Alzheimer’s disease (AD)- and Parkinson’s disease (PD)-related mortality among U.S. adults and to assess whether racial mortality patterns and temporal changes differed.

**Methods:** We analyzed CDC WONDER Multiple Cause of Death data from 2018-2024, where mortality was defined as deaths with AD or PD recorded as an underlying or contributing cause. Age-standardized mortality rates and adjusted mortality rate ratios (MRRs) were estimated. Joint-point regression evaluated racial differences, annual percentage changes (APCs), and disease-race-year interactions. Sensitivity analyses assessed the robustness of the findings.

**Results:** Black had significantly lower PD-related mortality than White (adjusted mortality rate ratios [MRRs]: 0.62-0.69; all p<0.001), whereas AD-related mortality showed no significant racial differences (MRRs: 0.91-1.06). Compared with 2018, mortality increased significantly in 2020 for both diseases among White adults (AD: MRR=1.21; PD: MRR=1.27) and Black adults (AD: MRR=1.27; PD: MRR=1.37), followed by partial declines. PD-related mortality increased significantly among White adults during 2018-2023 (annual percentage change [APC]=3.08%; 95% CI: 1.15%-5.05%), whereas trends among Black adults (APC=3.51%; 95% CI: -2.94% to 10.39%) and AD-related mortality trends in both racial groups were not statistically significant. The disease-race-year interaction was not significant (p=0.942), indicating insufficient evidence that temporal mortality patterns differed jointly.

**Conclusions:** Recorded PD-related mortality was lower among Black than White adults, whereas AD-related mortality showed no statistically significant racial difference. These findings support disease-specific mortality surveillance and further investigation of potential differences in diagnostic recognition and death-certificate documentation.

## Introduction

Alzheimer’s disease (AD) and Parkinson’s disease (PD) are major neurodegenerative disorders contributing substantially to disability, healthcare utilization, and mortality in the United States. Their growing burden presents important public health challenges, particularly as the population ages. An estimated 7.4 million Americans aged 65 years or older are living with Alzheimer’s dementia in 2026, with substantial racial variation in disease prevalence [1,2]. Evidence suggests that older Black adults experience a greater burden of Alzheimer’s and related dementias than White adults, whereas PD studies have reported lower rates of diagnosis among Black populations [2,3]. Racial inequities in neurological healthcare access further complicate these patterns; Black adults with neurological conditions have been found to have lower utilization of outpatient specialist care than White adults [4]. These differences underscore the importance of distinguishing underlying disease burden from recorded disease-related mortality, particularly among adults aged 45-74 years, for whom neurodegenerative mortality patterns remain less extensively characterized.

Racial differences in neurodegenerative disease-related mortality may be conceptualized within a multilevel framework encompassing disease occurrence, diagnostic recognition, access to specialized care, clinical progression, and death certification. These interconnected factors may contribute to variation in recorded mortality, which does not necessarily reflect differences in underlying disease burden or survival. Previous studies have documented racial disparities in PD diagnosis, treatment, and neurological healthcare utilization, particularly among Black adults [3–5]. From a methodological perspective, death-certificate-based mortality estimates may reflect variation in disease ascertainment and certification practices, in addition to underlying population-level disease patterns [6,7]. However, existing research has largely examined AD and PD separately, leaving limited comparative evidence on whether racial mortality patterns and temporal trends differ between these conditions. A unified, age-adjusted analytical framework incorporating disease–race–time interactions offers an opportunity to characterize these differences while acknowledging that population-level mortality data cannot establish their underlying mechanisms.

Despite growing evidence of racial differences in AD and PD, limited research has directly compared Black African American - White disparities in mortality involving these conditions or examined how these disparities have changed over time. Mortality involving AD or PD, defined as deaths in which either disease is recorded as an underlying or contributing cause on the death certificate, remains insufficiently characterized among adults younger than 75 years. To address these gaps, this study examines racial differences and temporal trends in AD- and PD-related mortality among Black/African American and White U.S. adults aged 45-74 years. By comparing both diseases within a common epidemiological framework, this study aims to characterize disease-specific racial mortality patterns and their temporal variation, providing evidence to inform neurodegenerative disease mortality surveillance.

## Methods

### Study Design and Data Source

This retrospective, population-based study examined temporal trends and racial disparities in AD- and PD-related mortality using the CDC WONDER Multiple Cause of Death database for 2018–2024 [8]. The database provides death certificate-derived mortality counts by demographic characteristics, calendar year, and International Classification of Diseases, Tenth Revision (ICD-10) codes, with corresponding population denominators.

## Study Population and Data Extraction

Eligible records included deaths occurring between 2018 and 2024 among individuals with AD (ICD-10 codes G30.0, G30.1, or G30.9) or PD (ICD-10 code G20) documented as an underlying or contributing cause of death. Records outside the study period, specified age range, racial groups, or disease definitions were excluded. Eligible records were aggregated by calendar year, race, 10-year age group, and disease. The eligibility criteria, sequential exclusions, and final analytic samples are presented in **Figure 1**.

**Figure 1.**
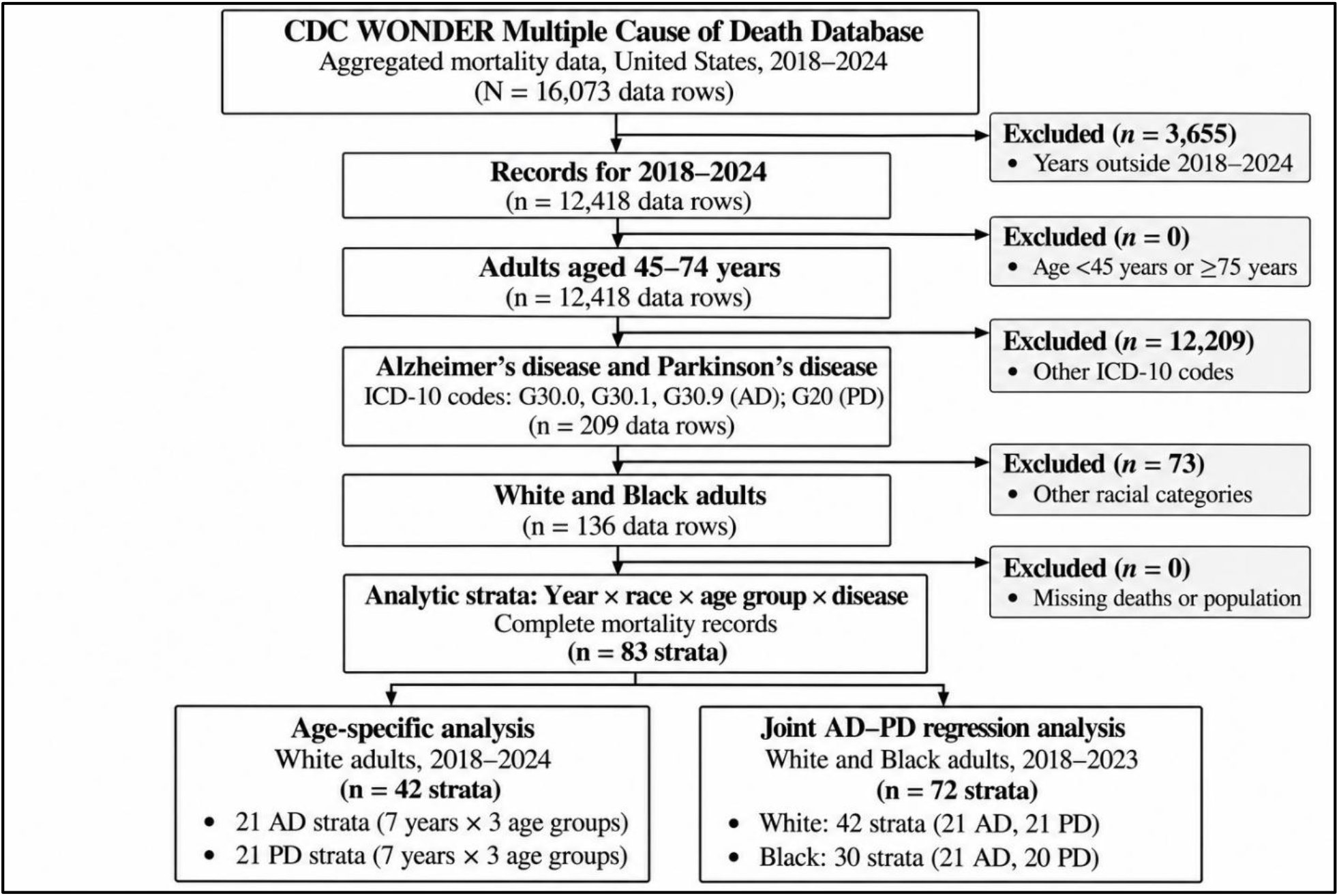
Data Extraction and Analytic Sample Selection for Alzheimer’s Disease-and Parkinson’s Disease-Related Mortality, CDC WONDER, 2018–2024

### Mortality Rate Calculation and Age Standardization

Annual AD- and PD-related mortality rates were calculated as the number of disease-related deaths divided by the corresponding population, multiplied by 100,000. Age-specific rates were estimated for adults aged 45–54, 55–64, and 65–74 years. Age-standardized mortality rates were calculated using direct standardization to account for differences in age distributions across racial groups and calendar years. Mortality rates were reported per 100,000 population with corresponding 95% confidence intervals.

### Statistical Analysis

Annual AD- and PD-related mortality rates were summarized by race, age group, and calendar year. Joint-point regression models with a log link and population offset were fitted to evaluate temporal trends and Black-White mortality disparities. The model was specified as:

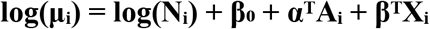

where μᵢ represents the expected death count, Nᵢ the population denominator, Aᵢ the age-group indicators, and Xᵢ the main effects and all two- and three-way interactions among disease, race, and calendar year. Overdispersion was accommodated using Var(Yᵢ) = φμᵢ, where φ denotes the dispersion parameter.

Calendar year was modeled categorically to estimate adjusted mortality rate ratios (RRs) and continuously to assess linear temporal trends. Disease- and race-specific annual percentage changes (APCs) were calculated as:

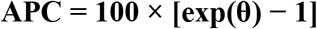

where θ represents the corresponding annual change in log mortality rate. Separate models incorporating disease-by-age-by-year interactions assessed age-specific trends among White adults. Joint racial analyses covered 2018–2023, whereas White age-specific analyses included 2018–2024.

Sensitivity analyses assessed the robustness of disease- and race-specific mortality trends using negative binomial regression as an alternative to joint-point modeling. Additional models excluded 2020 to evaluate the influence of the COVID-19 pandemic and separately examined pre-pandemic trends (2018–2019). Annual percentage changes (APCs) and corresponding 95% confidence intervals were compared across models to assess the consistency of trend direction, magnitude, and statistical significance. Adjusted mortality rates, RRs, and APCs were reported with 95% confidence intervals. Statistical significance was defined as a two-sided P < 0.05. This study used publicly available, aggregated, deidentified mortality data from CDC WONDER. Because no individual-level identifiable information was accessed, institutional review board approval and informed consent were not required.

## Results

### Analytic Sample and Descriptive Mortality Patterns

The final analytic dataset comprised 83 aggregated mortality strata, including 21 each for AD among White and Black adults and PD among White adults, and 20 for PD among Black adults (**Figure 1**). Age-standardized mortality rates increased across both diseases and racial groups in 2020 (**Table 1**; **Figure 2**). Among White adults, AD-related mortality increased from 8.60 (95% CI: 8.43–8.78) per 100,000 in 2018 to 10.41 (95% CI: 10.22–10.59) in 2020, while PD-related mortality increased from 7.17 (95% CI: 7.01–7.33) to 9.11 (95% CI: 8.94–9.29). Among Black adults, AD-related mortality increased from 8.74 (95% CI: 8.25–9.24) to 11.08 (95% CI: 10.54– 11.61), while PD-related mortality increased from 4.62 (95% CI: 4.26–4.98) to 6.33 (95% CI: 5.93–6.73). By 2024, AD- and PD-related mortality rates among White adults were 9.39 (95% CI: 9.22–9.57) and 8.19 (95% CI: 8.03–8.35) per 100,000, respectively, compared with 8.65 (95% CI: 8.21–9.10) for AD among Black adults.

**Figure 2.**
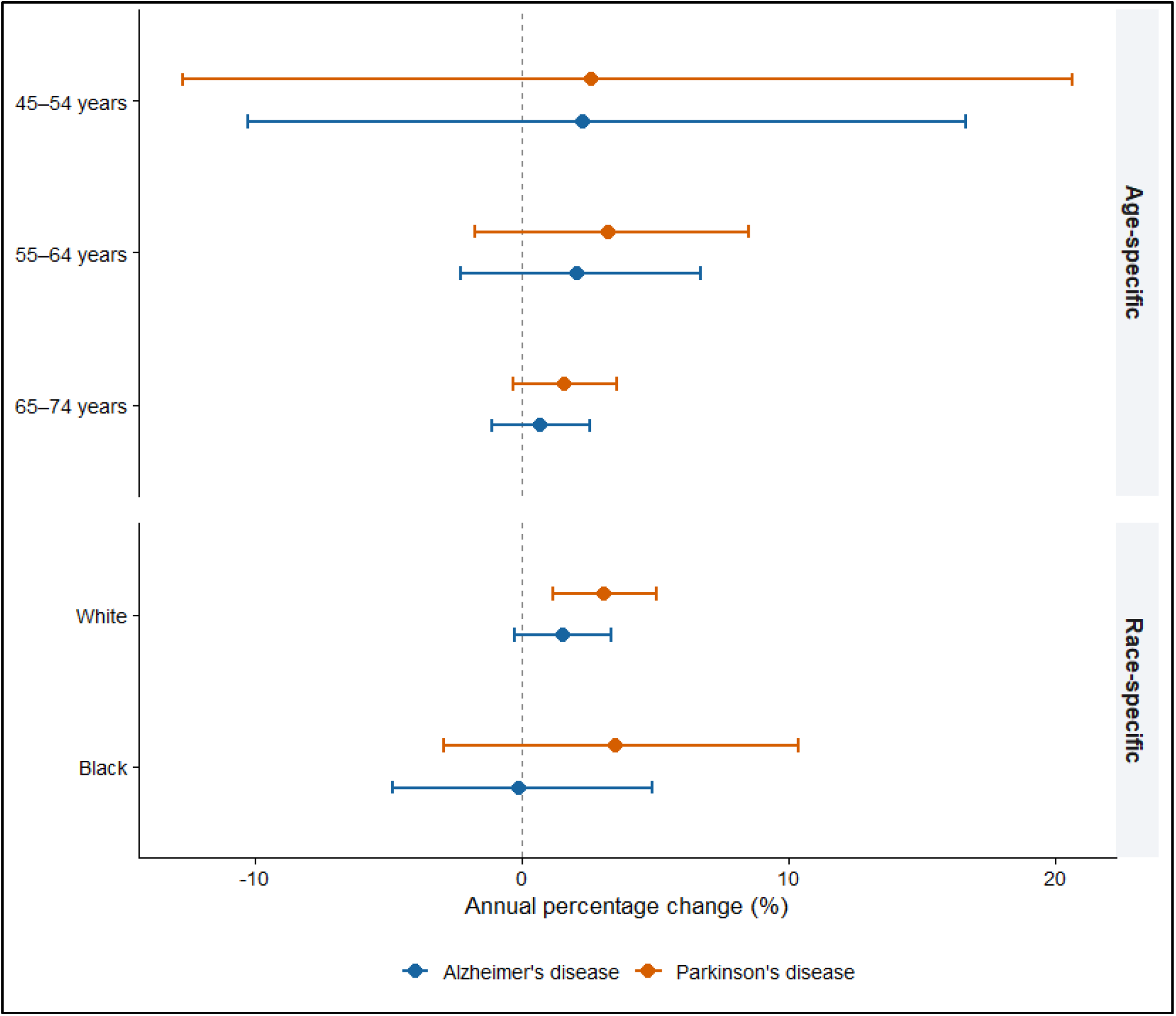
Annual Age-Standardized Alzheimer’s Disease-and Parkinson’s Disease-Related Mortality Rates by Race Among U.S. Adults Aged 45–74 Years, 2018–2024

**Table 1.** Annual Age-Standardized Parkinson’s Disease- and Alzheimer’s Disease-Related Mortality Rates by Race Among U.S. Adults Aged 45–74 Years, 2018–2024.

| Year | Race | Alzheimer's disease | Parkinson's disease |
| --- | --- | --- | --- |
| 2018 | White | 8.60 (8.43–8.78) | 7.17 (7.01–7.33) |
| 2018 | Black/ African American | 8.74 (8.25–9.24) | 4.62 (4.26–4.98) |
| 2019 | White | 8.76 (8.59–8.94) | 7.35 (7.19–7.51) |
| 2019 | Black/ African American | 8.72 (8.23–9.20) | 4.58 (4.23–4.94) |
| 2020 | White | 10.41 (10.22–10.59) | 9.11 (8.94–9.29) |
| 2020 | Black/ African American | 11.08 (10.54–11.61) | 6.33 (5.93–6.73) |
| 2021 | White | 9.56 (9.39–9.74) | 8.71 (8.54–8.88) |
| 2021 | Black/ African American | 10.01 (9.51–10.51) | 5.69 (5.32–6.07) |
| 2022 | White | 9.79 (9.61–9.97) | 8.84 (8.67–9.01) |
| 2022 | Black/ African American | 9.61 (9.13–10.10) | 5.86 (5.48–6.23) |
| 2023 | White | 9.22 (9.05–9.39) | 8.16 (8.00–8.32) |
| 2023 | Black/ African American | 8.37 (7.93–8.81) | 5.40 (5.04–5.75) |
| 2024 | White | 9.39 (9.22–9.57) | 8.19 (8.03–8.35) |
| 2024 | Black/ African American | 8.65 (8.21–9.10) | — |

Note: Rates per 100,000 population (95% confidence interval). Values are reproduced from your supplied table, not independently recalculated or validated. All other racial categories and unavailable rows have been removed. The 2024 Black PD estimate remains unavailable because the 45–54 age group is missing. AD estimates also require confirmation that ICD-10 subcode counts were not double-counted. Data limitation: AD code aggregation and potential overlap between AD- and PD-related death records remain unverified; inferential findings are provisional.

### Age-Adjusted Temporal Changes in AD- and PD-Related Mortality

Age-adjusted joint-point regression analyses demonstrated significant increases in AD- and PD-related mortality in 2020 relative to 2018 across both racial groups (**Table 2**). Among White adults, AD-related mortality increased by 21% (adjusted mortality rate ratio [MRR]: 1.21; 95% CI: 1.14–1.29; P < 0.001), while PD-related mortality increased by 27% (MRR: 1.27; 95% CI: 1.19–1.37; P < 0.001). Corresponding increases among Black adults were 27% for AD (MRR: 1.27; 95% CI: 1.06–1.52; P = 0.009) and 37% for PD (MRR: 1.37; 95% CI: 1.08–1.75; P = 0.011). Among White adults, mortality remained significantly elevated in 2023 for both AD (MRR: 1.07; 95% CI: 1.00–1.14; P = 0.049) and PD (MRR: 1.14; 95% CI: 1.06–1.22; P < 0.001), whereas corresponding changes among Black adults were not statistically significant. Across 2018–2023, PD-related mortality increased significantly among White adults (annual percentage change [APC]: 3.08%; 95% CI: 1.15%–5.05%; P = 0.002), while the AD trend was not statistically significant (APC: 1.52%; 95% CI: −0.27% to 3.33%; P = 0.096). Neither AD nor PD showed statistically significant linear trends among Black adults (AD APC: −0.13%; 95% CI: −4.88% to 4.87%; P = 0.959; PD APC: 3.51%; 95% CI: −2.94% to 10.39%; P = 0.293).

**Table 2.**
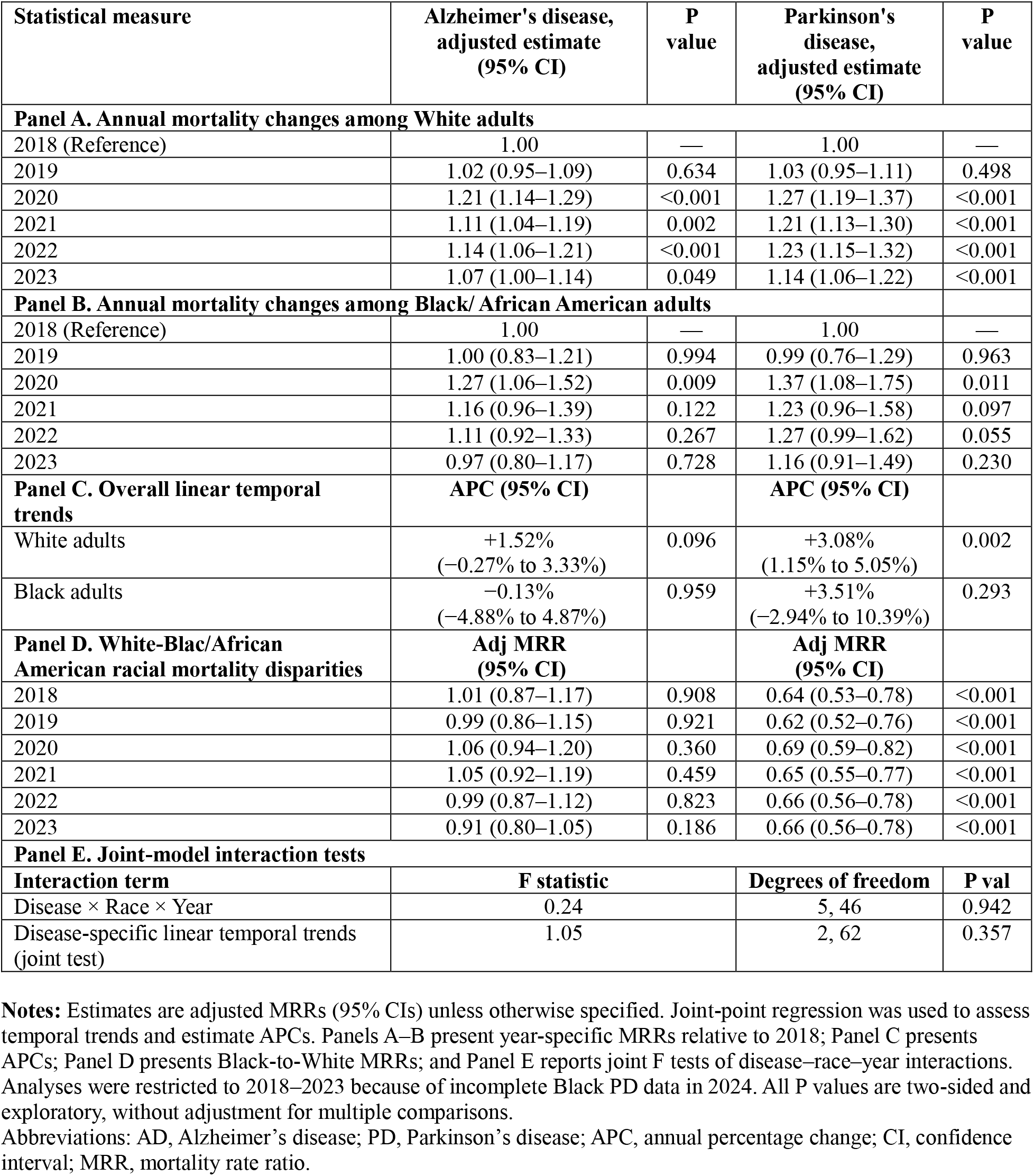
Age-Adjusted Temporal Changes and Racial Disparities in Alzheimer’s Disease-and Parkinson’s Disease-Related Mortality Among U.S. Adults Aged 45–74 Years, 2018–2023.

### Racial Disparities in AD- and PD-Related Mortality

Age-adjusted analyses revealed distinct racial patterns in AD- and PD-related mortality during 2018–2023 (Table 2). Black adults had consistently lower PD-related mortality than White adults, with adjusted Black-to-White mortality rate ratios (MRRs) ranging from 0.62 to 0.69 across study years (all P < 0.001). In 2018, Black adults had 36% lower PD-related mortality (MRR: 0.64; 95% CI: 0.53–0.78), and this disparity persisted in 2023 (MRR: 0.66; 95% CI: 0.56–0.78). In contrast, AD-related mortality did not differ significantly between Black and White adults in any study year, with adjusted MRRs ranging from 0.91 to 1.06 (all P > 0.05). In 2020, the adjusted Black-to-White MRR was 1.06 (95% CI: 0.94–1.20; P = 0.360) for AD compared with 0.69 (95% CI: 0.59–0.82; P < 0.001) for PD. The disease × race × year interaction was not statistically significant (F[5, 46] = 0.24; P = 0.942), providing no evidence that the joint disease-specific racial disparity patterns varied across calendar years.

### 3.4. Age-Specific Trends in AD- and PD-Related Mortality

Age-specific analyses among White adults during 2018–2024 showed positive but statistically nonsignificant annual percentage changes (APCs) in Alzheimer’s disease (AD)- and Parkinson’s disease (PD)-related mortality across all three age groups (Table 3, Panel A). Among adults aged 45–54 years, the APC was 2.29% (95% CI: −10.30% to 16.65%; P = 0.735) for AD and 2.61% (95% CI: −12.73% to 20.65%; P = 0.755) for PD. Corresponding APCs among adults aged 55– 64 years were 2.09% (95% CI: −2.30% to 6.68%; P = 0.356) for AD and 3.24% (95% CI: −1.79% to 8.53%; P = 0.210) for PD. Among adults aged 65–74 years, APCs were 0.70% (95% CI: −1.12% to 2.56%; P = 0.451) for AD and 1.59% (95% CI: −0.35% to 3.56%; P = 0.109) for PD. Age-by-year interactions were not statistically significant for either AD (P = 0.810) or PD (P = 0.858), indicating no evidence of differences in linear mortality trends across age groups.

**Table 3.**
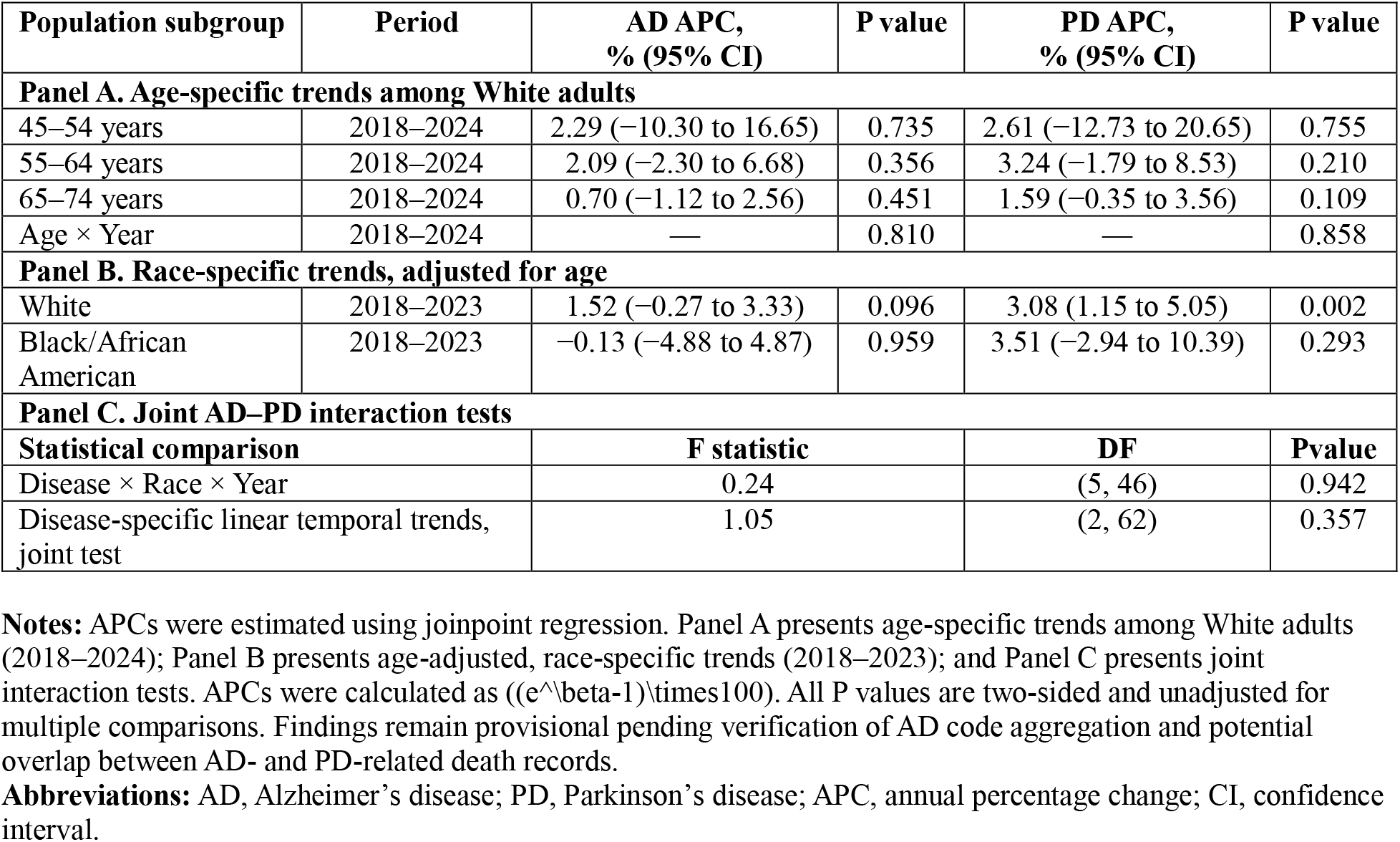
Age- and Race-Specific Annual Percentage Changes in Alzheimer’s Disease-and Parkinson’s Disease-Related Mortality Among U.S. Adults Aged 45–74 Years, 2018–2024.

### 3.5. Joint Disease-Specific Temporal Trend Comparisons

Joint-point regression analyses showed no statistically significant differences in temporal mortality patterns between Alzheimer’s disease (AD) and Parkinson’s disease (PD) across racial groups during 2018–2023 (Table 2, Panel E; Table 3, Panel C). The disease × race × year interaction was not significant (F[5, 46] = 0.24; P = 0.942), indicating insufficient evidence of differential year-specific mortality patterns by disease and race. Similarly, the joint test of disease-specific linear temporal trends was not significant (F[2, 62] = 1.05; P = 0.357), despite a statistically significant increase in PD-related mortality among White adults. These findings indicate that the observed differences in subgroup-specific annual percentage changes did not translate into statistically significant differences in the corresponding joint trend comparisons.

### 3.6. Model-Adjusted AD- and PD-Related Mortality Rates

Model-adjusted mortality rates demonstrated distinct disease-specific racial patterns during 2018–2023 (Figure 3; Supplementary Table S1). Among White adults, Alzheimer’s disease (AD)- related mortality increased from 3.80 (95% CI: 3.58–4.04) per 100,000 in 2018 to 4.60 (95% CI: 4.35–4.87) in 2020, before declining to 4.06 (95% CI: 3.83–4.30) in 2023. Corresponding rates among Black adults were 3.83 (95% CI: 3.33–4.42), 4.88 (95% CI: 4.32–5.51), and 3.71 (95% CI: 3.25–4.23), respectively. For Parkinson’s disease (PD), adjusted mortality rates among White adults increased from 3.17 (95% CI: 2.98–3.39) in 2018 to 4.04 (95% CI: 3.81–4.29) in 2020, followed by 3.61 (95% CI: 3.40–3.84) in 2023. Among Black adults, corresponding PD-related mortality rates were consistently lower, at 2.05 (95% CI: 1.69–2.48), 2.80 (95% CI: 2.39–3.29), and 2.38 (95% CI: 2.02–2.80), respectively. Both diseases exhibited their highest model-adjusted mortality rates in 2020 across racial groups, with more pronounced and persistent Black–White differences observed for PD than AD.

**Figure 3.**
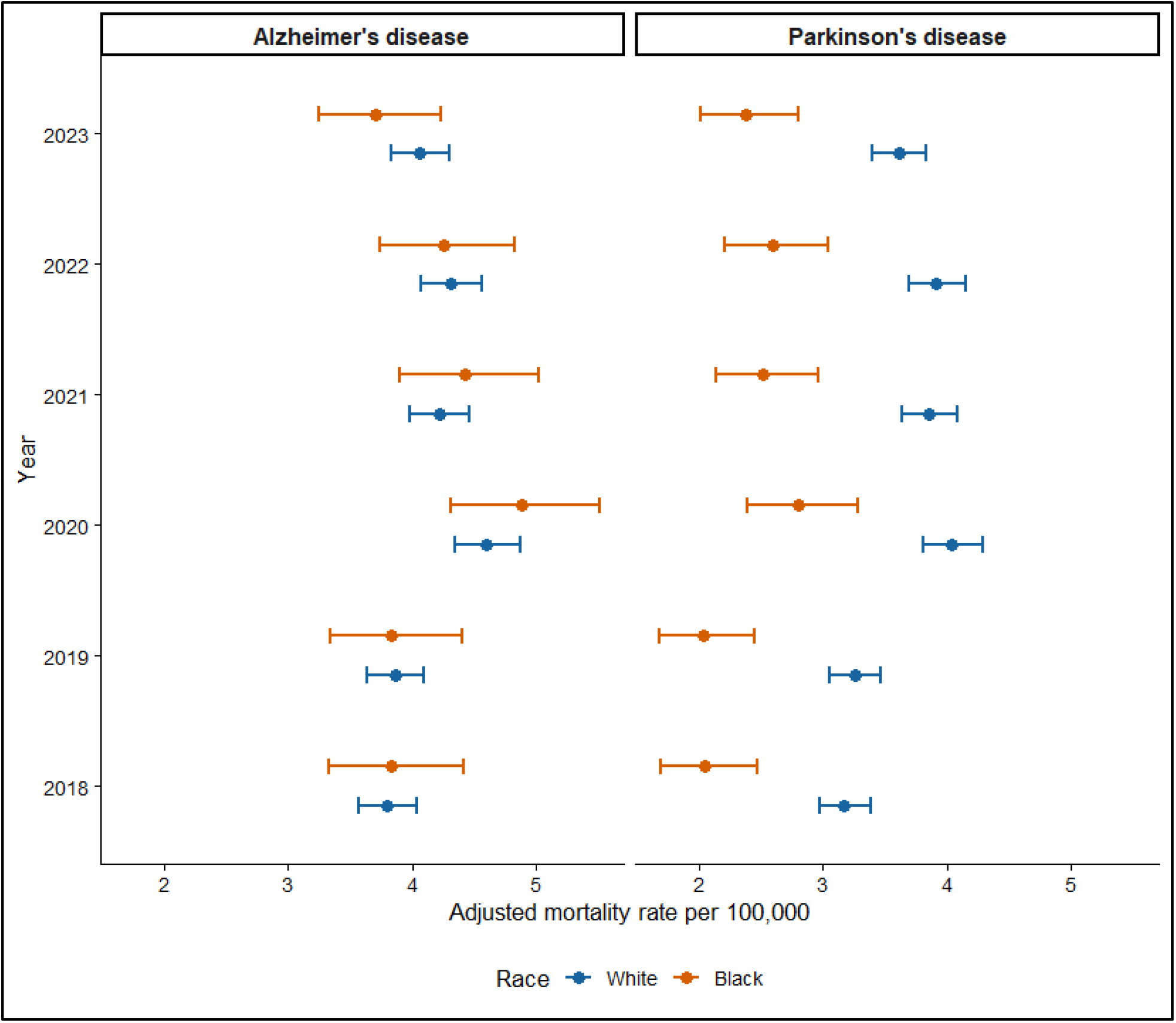
Model-Adjusted Alzheimer’s Disease- and Parkinson’s Disease-Related Mortality Rates by Race, 2018– 2023

### Sensitivity Analyses

Sensitivity analyses generally supported the direction of the primary mortality trends, although estimates varied across model specifications (Supplementary Table S3). In negative binomial regression, Parkinson’s disease (PD)-related mortality increased significantly among White adults (APC: 4.02%; 95% CI: 0.78%–7.37%; P = 0.015) and Black adults (APC: 4.48%; 95% CI: 0.46%–8.65%; P = 0.029), whereas Alzheimer’s disease (AD) trends remained nonsignificant. After excluding 2020, significant annual increases persisted among White adults for both AD (APC: 2.04%; 95% CI: 0.63%–3.47%; P = 0.004) and PD (APC: 3.68%; 95% CI: 2.15%–5.24%; P < 0.001), while trends among Black adults remained nonsignificant. Analyses restricted to the pre-pandemic period (2018–2019) yielded no statistically significant trends for either disease or racial group, with wide confidence intervals reflecting the limited observation period. Overall, sensitivity analyses supported increasing PD-related mortality among White adults, while evidence for trends in other subgroups was less consistent.

## Discussion

This nationwide population-based study provides comparative evidence of racial disparities in AD- and PD-related mortality among U.S. adults aged 45-74 years. The principal finding was a marked disease-specific contrast: Black adults had consistently lower recorded PD-related mortality than White adults, whereas AD-related mortality showed no statistically significant racial differences. Both diseases exhibited elevated mortality in 2020, followed by partial declines, and PD-related mortality increased significantly among White adults during 2018– 2023. However, joint interaction analyses did not establish statistically significant differences in temporal trends across diseases and racial groups. Collectively, these findings demonstrate the value of examining racial mortality disparities separately by neurodegenerative disease rather than assuming a uniform pattern across conditions. Importantly, the observed differences concern mortality recorded on death certificates and should not be interpreted as direct evidence of differences in underlying disease burden or survival.

The observed racial mortality patterns may be interpreted through a multilevel framework encompassing disease occurrence, clinical recognition, access to neurological care, disease progression, and death certification. Within this framework, recorded disease-related mortality may reflect both underlying disease patterns and variation in clinical recognition and death-certificate documentation [6,9]. Previous studies have demonstrated racial disparities in neurological healthcare utilization, including reduced access to specialist care among Black adults and differences in PD management and clinical outcomes [4,5]. These disparities may influence diagnostic ascertainment and subsequent documentation of PD on death certificates.

Furthermore, documented racial differences in dementia classification on death certificates suggest that recorded AD-related mortality may be influenced by variation in diagnostic classification and disease attribution [10]. Taken together, these findings underscore the importance of interpreting racial differences in recorded AD-and PD-related mortality within the broader context of disease ascertainment, healthcare access, and death-certification practices, rather than assuming that they directly reflect differences in disease occurrence or survival.

The pronounced increase in AD-and PD-related mortality in 2020, followed by partial declines, coincided with the COVID-19 pandemic and is consistent with previously reported excess mortality involving both conditions [11]. Potential contributing factors include vulnerability to severe infection and disruptions in neurological care, rehabilitation, and chronic disease management, although their relative contributions cannot be determined from the present data [12]. Previous studies have reported excess mortality among individuals with dementia and disruptions in care among patients with PD during the COVID-19 pandemic, suggesting that both infection-related vulnerability and changes in healthcare delivery may have contributed to observed mortality patterns [12–14]. Importantly, the persistence of elevated PD-related mortality among White adults beyond 2020 suggests that the observed temporal pattern was not confined to the initial pandemic year. However, the absence of statistically significant disease-specific differences in joint temporal trends and the ecological nature of the data preclude attributing these changes directly to COVID-19. These findings underscore the importance of examining pandemic-era mortality within the broader context of neurodegenerative disease progression, healthcare disruption, and changing mortality ascertainment.

Sensitivity analyses generally supported an increasing trend in PD-related mortality, particularly among White adults, although estimates varied across model specifications. Positive trends persisted after excluding 2020, suggesting that the increase was not attributable solely to the initial pandemic year. However, differences between joint-point and negative binomial estimates, especially among Black adults, warrant caution in interpreting subgroup-specific trends. These findings underscore the importance of race-and age-stratified mortality surveillance that considers potential differences in diagnostic recognition, access to neurological care, and death-certificate documentation [3–6]. Improving diagnostic access and mortality ascertainment may facilitate more accurate assessments of disparities, particularly because lower recorded PD-related mortality among Black adults does not necessarily indicate lower disease burden or improved survival.

This study has several strengths, including the use of nationwide mortality data, a comparative analytical framework for AD and PD, age-adjusted evaluation of racial disparities, and sensitivity analyses using alternative regression specifications. Nevertheless, several limitations warrant consideration. First, death certificate-based ascertainment is susceptible to diagnostic misclassification, underreporting, and racial differences in disease recognition and certification practices. Second, aggregated data precluded adjustment for individual-level socioeconomic characteristics, comorbidities, disease severity, healthcare utilization, and other potential confounders. Third, AD mortality counts derived from multiple ICD-10 subcodes may include duplicate decedents, and potential overlap between AD-and PD-related deaths was not explicitly accommodated in the joint regression models, potentially affecting estimates and statistical inference. Fourth, incomplete age-specific PD data among Black adults in 2024 restricted joint racial analyses to 2018–2023. Finally, the limited number of annual observations, unmodeled temporal dependence, and multiple exploratory comparisons may affect the precision and reliability of trend estimates. Accordingly, findings should be interpreted as descriptive population-level associations rather than evidence of causal relationships or underlying racial differences in disease incidence or survival.

## Conclusions

This nationwide study identified differing racial patterns in AD- and PD-related mortality among U.S. adults aged 45-74 years. Black adults had lower recorded PD-related mortality than White adults, whereas AD-related mortality showed no statistically significant racial differences. Both diseases exhibited elevated mortality in 2020, and PD-related mortality increased among White adults during 2018-2023, although differences in temporal trends across diseases and racial groups were not statistically significant. These findings underscore the value of disease-specific, race-stratified mortality surveillance and warrant further investigation into factors potentially associated with the observed disparities.

## Data Availability

CDC WONDER website

https://wonder.cdc.gov/

## Acknowledgments

The author gratefully acknowledges the University of South Florida for providing institutional start-up research support.

## Author Contributions

Md Roungu Ahmmad: Conceptualization, Methodology, Software, Validation, Formal analysis, Investigation, Resources, Data curation, Writing – original draft, Writing – review & editing, Visualization, Project administration, and Funding acquisition.

---

**eTable S1.**
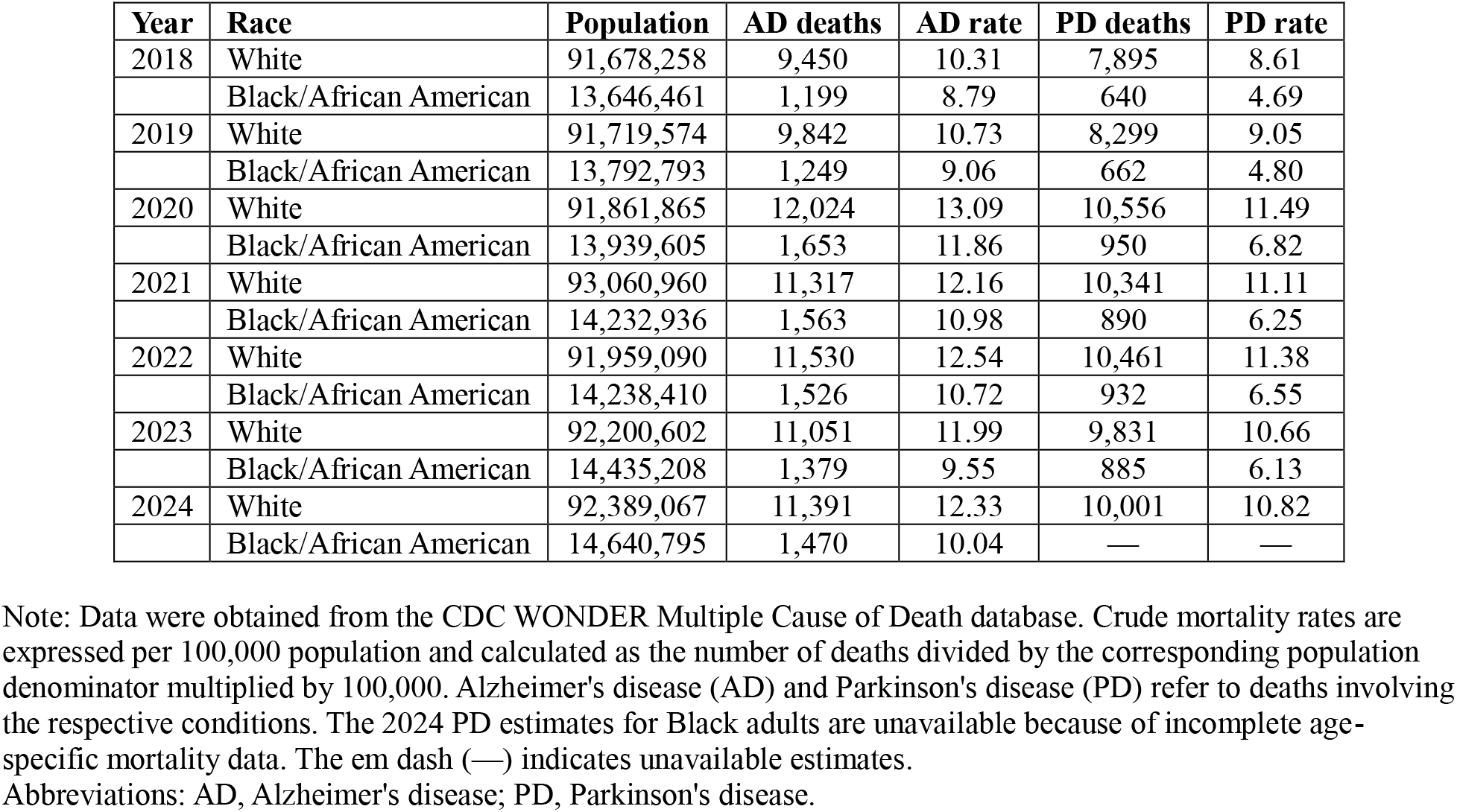
Annual Alzheimer’s Disease-and Parkinson’s Disease-Related Death Counts and Crude Mortality Rates by Race Among U.S. Adults Aged 45–74 Years, 2018–2024.

**eTable S2.**
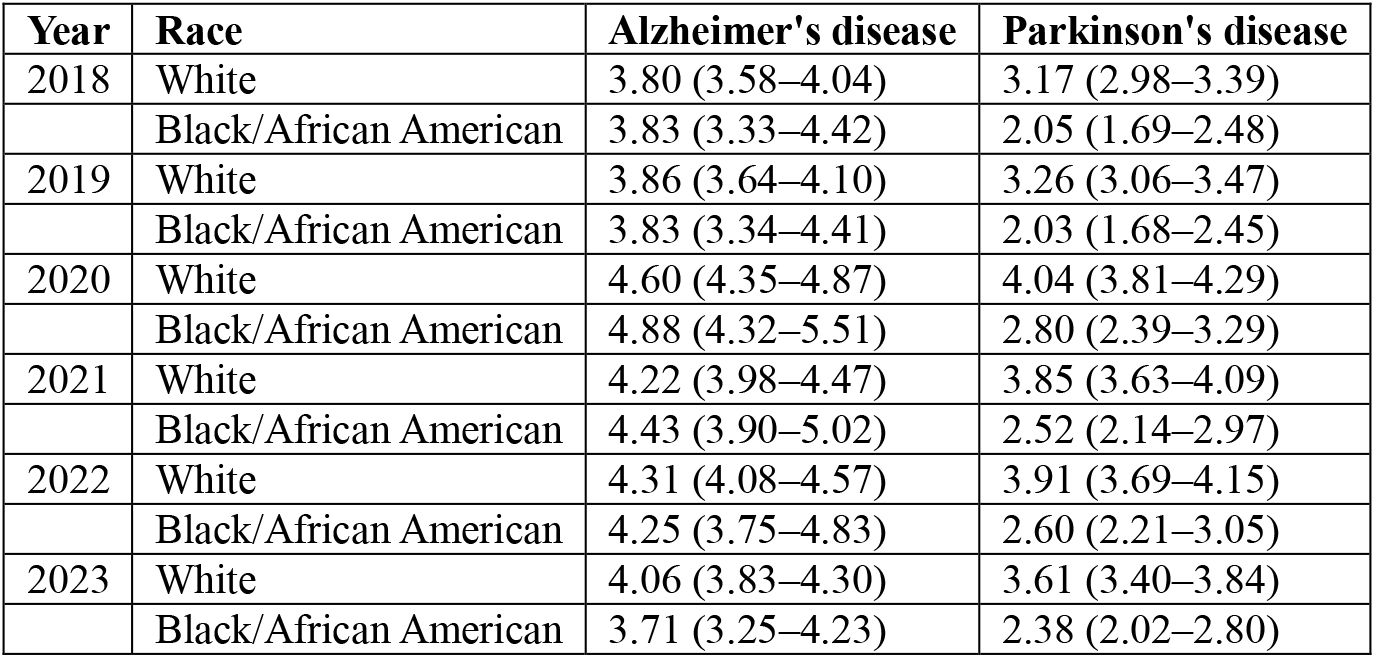
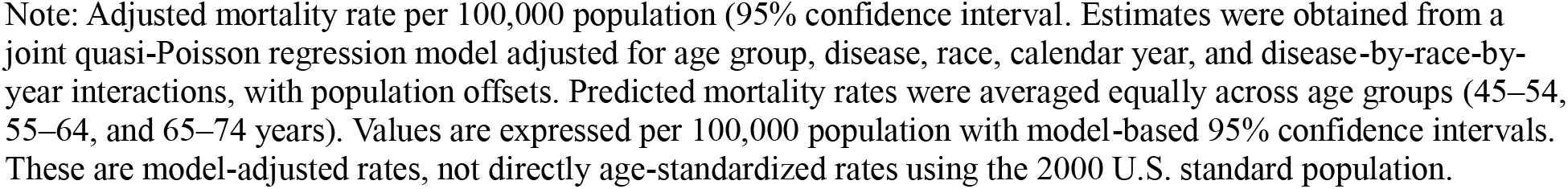
Model-Adjusted Alzheimer’s Disease- and Parkinson’s Disease-Related Mortality Rates by Race Among U.S. Adults Aged 45–74 Years, 2018–2023.

| Year | Race | Alzheimer's disease | Parkinson's disease |
| --- | --- | --- | --- |
| 2018 | White | 3.80 (3.58–4.04) | 3.17 (2.98–3.39) |
|  | Black/African American | 3.83 (3.33–4.42) | 2.05 (1.69–2.48) |
| 2019 | White | 3.86 (3.64–4.10) | 3.26 (3.06–3.47) |
|  | Black/African American | 3.83 (3.34–4.41) | 2.03 (1.68–2.45) |
| 2020 | White | 4.60 (4.35–4.87) | 4.04 (3.81–4.29) |
|  | Black/African American | 4.88 (4.32–5.51) | 2.80 (2.39–3.29) |
| 2021 | White | 4.22 (3.98–4.47) | 3.85 (3.63–4.09) |
|  | Black/African American | 4.43 (3.90–5.02) | 2.52 (2.14–2.97) |
| 2022 | White | 4.31 (4.08–4.57) | 3.91 (3.69–4.15) |
|  | Black/African American | 4.25 (3.75–4.83) | 2.60 (2.21–3.05) |
| 2023 | White | 4.06 (3.83–4.30) | 3.61 (3.40–3.84) |
|  | Black/African American | 3.71 (3.25–4.23) | 2.38 (2.02–2.80) |

Note: Adjusted mortality rate per 100,000 population (95% confidence interval). Estimates were obtained from a joint quasi-Poisson regression model adjusted for age group, disease, race, calendar year, and disease-by-race-by-year interactions, with population offsets. Predicted mortality rates were averaged equally across age groups (45–54, 55–64, and 65–74 years). Values are expressed per 100,000 population with model-based 95% confidence intervals. These are model-adjusted rates, not directly age-standardized rates using the 2000 U.S. standard population.

**eTable S3.**
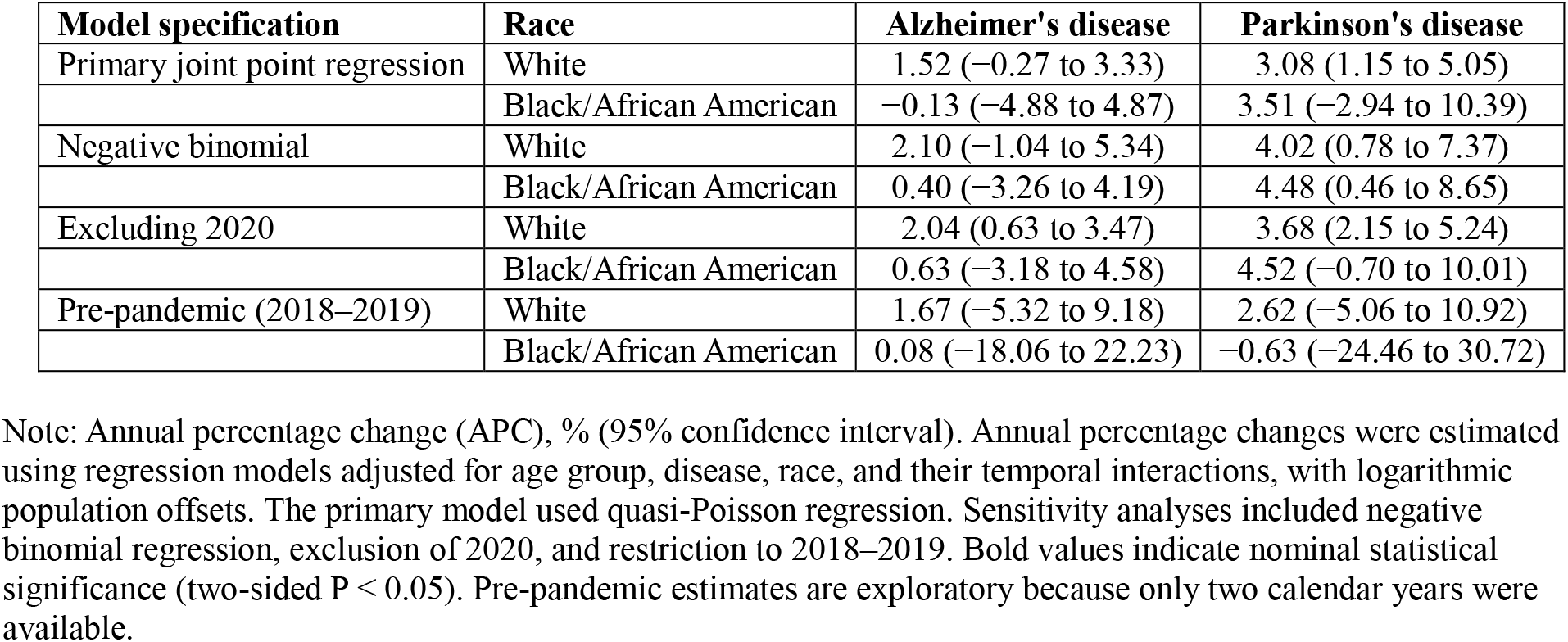
Sensitivity Analyses of Temporal Trends in Alzheimer’s Disease-and Parkinson’s Disease-Related Mortality Among U.S. Adults Aged 45–74 Years, 2018–2023.

| **Model specification** | **Race** | **Alzheimer's disease** | **Parkinson's disease** |
| --- | --- | --- | --- |
| Primary joint point regression | White | 1.52 (–0.27 to 3.33) | 3.08 (1.15 to 5.05) |
|  | Black/African American | –0.13 (–4.88 to 4.87) | 3.51 (–2.94 to 10.39) |
| Negative binomial | White | 2.10 (–1.04 to 5.34) | 4.02 (0.78 to 7.37) |
|  | Black/African American | 0.40 (–3.26 to 4.19) | 4.48 (0.46 to 8.65) |
| Excluding 2020 | White | 2.04 (0.63 to 3.47) | 3.68 (2.15 to 5.24) |
|  | Black/African American | 0.63 (–3.18 to 4.58) | 4.52 (–0.70 to 10.01) |
| Pre-pandemic (2018–2019) | White | 1.67 (–5.32 to 9.18) | 2.62 (–5.06 to 10.92) |
|  | Black/African American | 0.08 (–18.06 to 22.23) | –0.63 (–24.46 to 30.72) |

Note: Annual percentage change (APC), % (95% confidence interval). Annual percentage changes were estimated using regression models adjusted for age group, disease, race, and their temporal interactions, with logarithmic population offsets. The primary model used quasi-Poisson regression. Sensitivity analyses included negative binomial regression, exclusion of 2020, and restriction to 2018–2019. Bold values indicate nominal statistical significance (two-sided $P < 0.05$ ). Pre-pandemic estimates are exploratory because only two calendar years were available.

## References

1. Alzheimer’s Association [Internet]. [cited 2026 Oct 19]. Alzheimer’s Disease Facts and Figures. Available from: https://www.alz.org/alzheimers-dementia/facts-figures

2. Blank RH. Alzheimer’s Disease and Other Dementias: An Introduction. In: Blank RH, editor. Social & Public Policy of Alzheimer’s Disease in the United States [Internet]. Singapore: Springer; 2019 [cited 2025 Aug 22]. p. 1–26. Available from: 10.1007/978-981-13-0656-3_1 doi:10.1007/978-981-13-0656-3_1

3. Dahodwala N, Siderowf A, Xie M, Noll E, Stern M, Mandell DS. Racial Differences in the Diagnosis of Parkinson’s Disease. Mov Disord. 2009 Jun 15;24(8):1200–5. doi:10.1002/mds.22557 PubMed PMID: 19412929; PubMed Central PMCID: PMC2858583.

4. Saadi A, Himmelstein DU, Woolhandler S, Mejia NI. Racial disparities in neurologic health care access and utilization in the United States. Neurology. 2017 Jun 13;88(24):2268–75. doi:10.1212/WNL.0000000000004025 PubMed PMID: 28515272; PubMed Central PMCID: PMC5567325.

5. Willis AW, Schootman M, Evanoff BA, Perlmutter JS, Racette BA. Neurologist care in Parkinson disease: a utilization, outcomes, and survival study. Neurology. 2011 Aug 30;77(9):851–7. doi:10.1212/WNL.0b013e31822c9123 PubMed PMID: 21832214; PubMed Central PMCID: PMC3162639.

6. Benito-León J, Louis ED, Villarejo-Galende A, Romero JP, Bermejo-Pareja F. Under- reporting of Parkinson’s disease on death certificates: a population-based study (NEDICES). J Neurol Sci. 2014 Dec 15;347(1–2):188–92. doi:10.1016/j.jns.2014.08.048 PubMed PMID: 25292414.

7. Wolters FJ, Harteloh PPM, van Rooij FJA, Bakx P, van Heemst J, Kieboom BCT, et al. Diagnostic value of death certificates for case ascertainment of dementia: A population-based assessment of mortality registry data. J Alzheimers Dis. 2026 Apr;110(3):1428–37. doi:10.1177/13872877261423958 PubMed PMID: 41742625; PubMed Central PMCID: PMC13022020.

8. Multiple Cause of Death 2018-2024 by Single Race [Internet]. [cited 2026 Oct 20]. Available from: https://wonder.cdc.gov/wonder/help/mcd-expanded.html?utm_source=chatgpt.com

9. Romero JP, Benito-León J, Mitchell AJ, Trincado R, Bermejo-Pareja F. Under Reporting of Dementia Deaths on Death Certificates using Data from A Population-Based Study (NEDICES). Journal of Alzheimer’s Disease. 2014 Feb 14;39(4):741–8. doi:10.3233/JAD-131622

10. Gillum RF, Obisesan TO. Differences in mortality associated with dementia in U.S. blacks and whites. J Am Geriatr Soc. 2011 Oct;59(10):1823–8. doi:10.1111/j.1532-5415.2011.03598.x PubMed PMID: 22091495; PubMed Central PMCID: PMC3228365.

11. Li S, Han L, Shi H, Chong MKC, Zhao S, Ran J. Excess deaths from Alzheimer’s disease and Parkinson’s disease during the COVID-19 pandemic in the USA. Age Ageing. 2022 Dec 5;51(12):afac277. doi:10.1093/ageing/afac277 PubMed PMID: 36571781.

12. Brown EG, Chahine LM, Goldman SM, Korell M, Mann E, Kinel DR, et al. The Effect of the COVID-19 Pandemic on People with Parkinson’s Disease. J Parkinsons Dis. 2020;10(4):1365–77. doi:10.3233/JPD-202249 PubMed PMID: 32925107; PubMed Central PMCID: PMC7683050.

13. Azarpazhooh MR, Morovatdar N, Avan A, Phan TG, Divani AA, Yassi N, et al. COVID-19 Pandemic and Burden of Non-Communicable Diseases: An Ecological Study on Data of 185 Countries. Journal of Stroke and Cerebrovascular Diseases. 2020 Sep 1;29(9). doi:10.1016/j.jstrokecerebrovasdis.2020.105089 PubMed PMID: 32807484.

14. Cagnin A, Di Lorenzo R, Marra C, Bonanni L, Cupidi C, Laganà V, et al. Behavioral and Psychological Effects of Coronavirus Disease-19 Quarantine in Patients With Dementia. Front Psychiatry. 2020 Sep 9;11. doi:10.3389/fpsyt.2020.578015

